# Rising dengue burden in high-altitude central Nepal: evidence from a population-based longitudinal serosurvey, 2019-2023

**DOI:** 10.64898/2026.07.14.26357903

**Authors:** Aastha Shrestha, Melina Thapa, Sony Shrestha, Sudichhya Tamrakar, Urursha Ranjitkar, Nishan Katuwal, Sabin Bikram Shahi, Shiva Ram Naga, Jason R. Andrews, Rajeev Shrestha, Kristen Aiemjoy, Dipesh Tamrakar

## Abstract

**Background:** Dengue is intensifying globally due to climate change, urbanization, and land use changes. In Nepal, dengue has expanded from lowland regions to higher altitudes, with record outbreaks in 2022 and 2023. However, reliance on passive surveillance and hospital-based studies may underestimate community-level infection burden.

**Methods:** We conducted a population-based serologic cohort study in Kathmandu and Kavrepalanchok districts, Nepal, enrolling a geographically representative, age-stratified random sample of residents aged 0–25 years from pre-defined hospital catchment areas. Enrollment occurred in two phases: Phase I (February 2019 to April 2021) with follow-up visits at approximately 3, 6, and 12 months, and Phase II (February to June 2023) revisiting the original cohort. At each household visit, we collected capillary blood samples by finger-prick onto filter paper and tested the samples for IgG responses against dengue-derived recombinant antigen using InBios DENV Detect^TM^ ELISA. Serostatus was classified using the manufacturer’s recommended immune status ratio (ISR) cutoffs. We calculated seroprevalence at each time point and estimated seroincidence rates by identifying seroconversion events per 1,000 person-years. We assessed risk factors using multivariable regression models.

**Results:** Between 2019 and 2023, we enrolled 840 participants and collected 2,082 blood samples. The overall seroincidence rate was 33.8 per 1,000 person-years (95% CI [24.9-45.0]), with the highest rates in urban Kathmandu ([105.7], 95% CI [75.1-144.4]). Seroincidence increased with age and over time from 46.1 in 2019 to 51.0 in 2023. Participants living with a dengue-positive individual in the same household (adjusted RR [4.65], 95% CI [2.72–8.0]) and households with water-filled flower basins (adjusted RR [2.53], 95% CI [1.28–5.74]) had significantly higher risk of seroconversion.

**Conclusions:** This study reveals a significant and increasing burden of dengue infection in the Kathmandu Valley between 2019 and 2023. highlighting an urgent need for immediate public health interventions to mitigate dengue’s rise in Nepal’s higher-altitude regions.

## BACKGROUND

Dengue is a widespread mosquito-borne viral infection transmitted by *Aedes* mosquitoes (1,2). There are an estimated 59 million infections and 29 thousand deaths annually; approximately 70% of infections occur in Asia (3). The global incidence has more than doubled every decade between 1990 and 2021, with South Asia, Southeast Asia, and tropical Latin America being the most severely affected regions (4). This recent rise in burden may be driven by climate change, urbanization and increased human mobility (5,6).

In Nepal, the first imported case of dengue was reported in 2004 and local transmission was documented in 2006 (7–9). Since then, sporadic outbreaks have occurred, typically in two to three-year cycles. Initially, dengue was limited to the lowland (Terai) regions of Nepal but cases have been increasingly reported in the hill and mountain regions since 2017, with unprecedented outbreaks at high altitudes (10,11). A large outbreak occurred in 2019 with over 17,000 reported cases followed by a record outbreak in 2022 affecting all 77 districts with over 54,000 reported cases (11–13). Molecular surveillance has identified co-circulation of multiple dengue virus serotypes, with DENV-1, DENV-2, and DENV-3 detected during the 2022 outbreak and DENV-2 predominating in 2023 (12,14). Phylogenetic analysis reveals that circulating strains are closely related to Indian and Southeast Asian lineages, indicating multiple introductions and ongoing viral evolution (7,14–16). The proportion of secondary infections—which carry increased risk of severe disease through antibody-dependent enhancement—varies by outbreak and geographic location, with 60% secondary infections documented during the 2010 outbreaks in central and western Nepal and 18.3% during the 2023 outbreak in eastern Nepal (4,9,14).

Dengue presents with a wide clinical spectrum, ranging from self-limiting febrile illness with fever, headache, myalgia, and retro-orbital pain to severe dengue characterized by plasma leakage, hemorrhagic manifestations, and shock. Most dengue surveillance systems are passive or facility-based, which can be biased by factors such as healthcare accessibility and health care seeking behaviour (15,17–21). Moreover, clinical surveillance substantially underestimates the true burden of the infection because approximately 60-80% of dengue infections are asymptomatic or subclinical and do not seek care(4). Population-based cohort studies have consistently demonstrated that dengue cases are greatly under-reported through official passive surveillance systems, suggesting that reported cases represent only a fraction of true infections (22).

Serosurveys play an important role in monitoring the population-level infection burden of dengue (23) and can augment clinical surveillance by detecting subclinical infections that would otherwise go undetected as well as clinical infections that are not diagnosed. Seroprevalence reflects cumulative exposure and is shaped by population age structure, underlying transmission intensity, and antibody kinetics after infection. Seroprevalence estimates vary widely across South Asia, with high IgG antibody seroprevalence (>60%) reported in highly endemic areas such as Bangladesh, Thailand, Indonesia, Sri Lanka and India and mid-range (10-60%) seroprevalence in areas with more sporadic transmission (24). In neighboring India, a nationally representative serosurvey conducted in 2017 found an overall dengue seroprevalence of 48.7%, with significant geographic heterogeneity and higher seroprevalence in southern (76.9%), western (62.3%), and northern (60.3%) regions (25). Longitudinal serosurveys permit estimation of the force of infection (FOI)—the per capita rate at which susceptible individuals become infected per unit time—which provides critical data on transmission dynamics in endemic populations. Studies estimating FOI in Asia have revealed substantial heterogeneity in transmission intensity. In India, site-specific FOI estimates ranged from 35 to 212 infections per 1000 person-years (26). Longitudinal serosurveys in Thailand documented seroconversion rates of 48 to 147 per 1000 person-years (27). Across 13 endemic countries in Asia, estimated FOI varied widely from 17 per 1000 person-year in Singapore to 241 per 1000 person-year in the Philippines (28,29). These FOI estimates demonstrate that dengue transmission intensity varies markedly both across and within countries and over time.

Prior studies of dengue transmission and exposure in Nepal have been limited to hospital settings or have investigated knowledge, attitude and behaviours at the population level (30,31). To our knowledge, there have been no prior community-based serosurveys of dengue in Nepal. Given the rapid geographic expansion of dengue to previously non-endemic areas, the increasing outbreak frequency and magnitude, the co-circulation of multiple serotypes with increasing secondary infections, and the substantial proportion of undetected infections, there is an urgent need to characterize the true burden of dengue infection in Nepal. In order to fill in this gap, we conducted representative population-based longitudinal cohort serologic study among the residents of Kathmandu and Kavre districts in Nepal. Our objectives were to characterize the infection burden of dengue at the community level, estimate seroprevalence and seroincidence, and identify the risk factors for infection to facilitate future dengue prevention and control efforts in Nepal.

## METHODS

### Participants enrollment

We enrolled the participants in two phases from pre-defined hospital catchment areas in Kathmandu and Kavrepalanchok districts. In **Phase I** (February 2019 to April 2021), we leveraged samples collected for an enteric fever serosurvey; details of the study design and sampling are described in detail in (32). In brief, we randomly selected an age-stratified sample of individuals aged 0–25 years from a previously conducted healthcare-seeking behavior survey. We visited participants at their households to obtain consent and enroll them into the study. After enrollment, we returned to their households for follow-up visits at approximately 3, 6, and 12 months to collect follow up blood samples. In **Phase II** (February to June 2023), we revisited households of participants from the original cohort who had agreed to participate in continued follow-up. We obtained informed consent (or assent with parental consent for minors) at the time of initial enrollment and reconsented during follow-up.

### Sample and data collection

During each household visit, we collected the capillary blood samples by finger-prick onto the filter paper (TropBio FP 05-002-12) to produce dried blood spot samples (DBS). We air-dried the DBS papers for at least 15 minutes, placed each sample in an individual plastic bag with desiccant, and stored them at -20 °C until testing.

We administered structured questionnaires using REDCap on tablets during both study phases. In Phase I, we collected demographic information only. In Phase II, we collected updated demographic data and assessed environmental and behavioral risk factors for dengue virus exposure. We recorded GPS coordinates at each household.

### Laboratory methods

We eluted the DBS by submerging a single blood filled filter protrusion in 66.6 µL of 1X Phosphate Buffer Saline (PBS) containing 0.05% Tween and incubated overnight at 4°C. The following day, we centrifuged the submerged sample at 10,000g for 5 mins and the supernatant was transferred in a new tube considering the dilution of DBS to be 1:10.

We tested the samples for IgG responses against dengue-derived recombinant antigen using a commercially available InBios DENV Detect^TM^ IgG ELISA kit (Cat: DDGS-R). Each sample was run in duplicate wells: one coated with dengue-derived recombinant antigen (DENRA) and one coated with normal cell antigen (NCA) to account for background signal. The assay was performed according to the manufacturer’s instructions, and optical density was measured at 450 nm using a Bio-Rad iMark ELISA plate reader. The assay output was expressed as the immune status ratio (ISR), calculated according to the manufacturer’s protocol using the ratio of antigen-specific signal to background signal relative to kit controls. The InBios kit was chosen based on published papers demonstrating a high diagnostic performance, with a sensitivity of 97% and specificity of 97.7% (33).

### Statistical methods

We classified dengue serostatus using ISR thresholds provided by the InBios DENV Detect™ kit. Samples were considered seropositive if ISR > 2.84, borderline if ISR 1.65–2.84, and seronegative if ISR ≤ 1.65. We calculated dengue seroprevalence as the proportion of individuals whose value was above the manufacturer-defined threshold at each time point, dividing the number of seropositive participants by the total number of individuals tested. We defined seroconversion as a change from seronegative to seropositive status between two consecutive visits, and seroreversion as a change from seropositive to seronegative over the same interval. To estimate seroincidence, we identified participants who seroconverted during follow-up and calculated person-time at risk using the midpoint between sample collection dates. We expressed seroincidence as the number of seroconversions divided by the total person-time at risk, assuming seroconversion occurred at the midpoint between the last negative and first positive samples. We used exact methods based on the Poisson distribution to calculate 95% confidence intervals for the incidence rate.

We used univariate log-binomial regression to identify potential risk factors associated with seroconversions. For each predictor variable, we fit a generalized linear model with a binomial distribution and a log link function to estimate the risk ratio (RR) and corresponding 95% confidence interval (CI).

As a sensitivity analysis, we repeated all analyses using an empirically derived seropositivity threshold to evaluate the impact of classification cutoffs on study conclusions. We fit a two-component Gaussian finite mixture model to the log-transformed ISR values from baseline samples. The alternative cutoff was defined as the mean of the lower mixture component plus three standard deviations on the log scale and then back-transformed to the original scale, yielding a cutoff value of ISR 1.55.

All analyses were performed using R version 4.3.3.

### Sample size

The sample size was determined by the parent enteric fever serosurvey, which enrolled an age-stratified random sample of 840 participants from Kathmandu and Kavrepalanchok districts. For dengue seroincidence estimation, approximately 1,400 person-years of follow-up provided reasonable precision for rates on the order of 25 per 1,000 person-years (about 35 expected seroconversion events). For the Phase II risk factor analysis (N=354 with complete risk factor data), power to detect associations depended on exposure prevalence and the underlying seroconversion risk; for exposures with prevalence ≥15%, we were well powered to detect large effects (e.g., risk ratios around 2 or greater when baseline seroconversion risk was moderate), but had limited power for smaller effects and uncommon exposures.

## RESULTS

### Socio-demographic characteristics

A total of 840 participants were enrolled from Kathmandu (N= 350) and Kavre (N=490) between 2019 and 2023. Participants provided 2082 blood samples over 1.0 - 5.0 study visits. The median age of participants was 11 (IQR 5.9-17), ranging from 1 to 28 years. 53.1% (446/840) of participants were male (Table 1).

**Table 1:** Summary of Study Population.

|  | **Kathmandu**<br>**(N=350)** | **Kavrepalanchok**<br>**(N=490)** | **Overall**<br>**(N=840)** |
| --- | --- | --- | --- |
| **Age, in years** |  |  |  |
| Median (IQR) | 13 (6.7 - 18) | 11 (5.4 - 17) | 11 (5.9 - 17) |
| Min/Max | 1.3 - 27 | 1.1 - 28 | 1.1 - 28 |
| **Gender** |  |  |  |
| Female | 164 (46.9%) | 230 (46.9%) | 394 (46.9%) |
| Male | 186 (53.1%) | 260 (53.1%) | 446 (53.1%) |
| **Number of study visits** |  |  |  |
| Median (IQR) | 2.0 (1.0 - 3.0) | 3.0 (2.0 - 4.0) | 2.0 (1.0 - 3.0) |
| Min/Max | 1.0 - 5.0 | 1.0 - 5.0 | 1.0 - 5.0 |
| **Current level of education** |  |  |  |
| Preschool | 49 (14.0%) | 107 (21.8%) | 156 (18.6%) |
| Primary school | 94 (26.9%) | 149 (30.4%) | 243 (28.9%) |
| Secondary school | 74 (21.1%) | 91 (18.6%) | 165 (19.6%) |
| College/University | 89 (25.4%) | 80 (16.3%) | 169 (20.1%) |
| Missing | 44 (12.6%) | 63 (12.9%) | 107 (12.7%) |

### Dengue seroprevalence by age, municipality and over time

The overall dengue seroprevalence was 4.7% (97/2082). Seroprevalence increased with age, rising from 2.6% (8/309) among 1-5 year olds to 7.7% (55/712) among 15-28 year olds. The seroprevalence between males and females were similar. Dengue seroprevalence increased over the study period, rising from 2.1% (16/761) in 2019 to 13.3% (47/354) in 2023. Geographically, the highest seroprevalence was observed in Kathmandu in 2023 at 34 % (36/106). In contrast, the seroprevalence in Dhulikhel was 1.8% (1/55) in 2023. (**Table 2**)

**Table 2:** Dengue Seroprevalence by year.

| var | 2019 | 2020 | 2021 | 2023 | Overall |
| --- | --- | --- | --- | --- | --- |
| Overall | 2.1% (16/761) | 4.3% (28/649) | 1.9% (6/318) | 13.3% (47/354) | 4.7% (97/2082) |
| Age, yrs |  |  |  |  |  |
| 1-<5 | 0.0% (0/149) | 7.0% (8/114) | 0.0% (0/43) | 0.0% (0/3) | 2.6% (8/309) |
| 5-<10 | 0.6% (1/169) | 2.0% (3/153) | 0.0% (0/82) | 7.7% (8/104) | 2.4% (12/508) |
| 10-<15 | 1.5% (3/201) | 1.2% (2/169) | 4.4% (4/90) | 14.0% (13/93) | 4.0% (22/553) |
| 15-28 | 5.0% (12/242) | 7.0% (15/213) | 1.9% (2/103) | 16.9% (26/154) | 7.7% (55/712) |
| City/town* |  |  |  |  |  |
| Banepa | 0.0% (0/170) | 2.8% (4/144) | 0.0% (0/64) | 5.3% (2/38) | 1.4% (6/416) |
| Dhulikhel | 0.0% (0/86) | 0.0% (0/105) | 0.0% (0/17) | 1.8% (1/55) | 0.4% (1/263) |
| Kathmandu | 4.3% (11/257) | 9.8% (22/224) | 5.9% (5/85) | 34.0% (36/106) | 11.0% (74/672) |
| Panauti | 2.7% (5/187) | 1.5% (2/134) | 0.9% (1/114) | 3.2% (4/124) | 2.1% (12/559) |
| Panchkhal | 0.0% (0/61) | 0.0% (0/42) | 0.0% (0/38) | 12.9% (4/31) | 2.3% (4/172) |
| Gender* |  |  |  |  |  |
| Female | 2.1% (8/374) | 4.8% (15/313) | 1.3% (2/151) | 13.6% (24/177) | 4.8% (49/1015) |
| Male | 2.1% (8/387) | 3.9% (13/336) | 2.4% (4/167) | 13.0% (23/177) | 4.5% (48/1067) |

### Dengue Seroincidence

The overall seroincidence rate was 33.8 per 1000 person-years (95% CI, 24.9-45.0) The seroincidence rose with age from 11.7 per 1000 person-years among the children of 0 to 5 years old to 52.0 per 1000 person-years among the participants 15-28 years old. Geographically, the seroincidence was lowest in Panauti with 2.2 per 1000 person-years (95% CI, 0.1-12.1) followed by Dhulikhel with 4.8 per 1000 person-years (95% CI, 0.1-27.0). Meanwhile, the highest seroincidence was observed in Kathmandu at 105.7 per 1000 person-years (95% CI, 75.1-144.4) (**Figure 2)**. There were no strong differences in the seroincidence rate by gender (**Table 3**).

**Figure 1.**
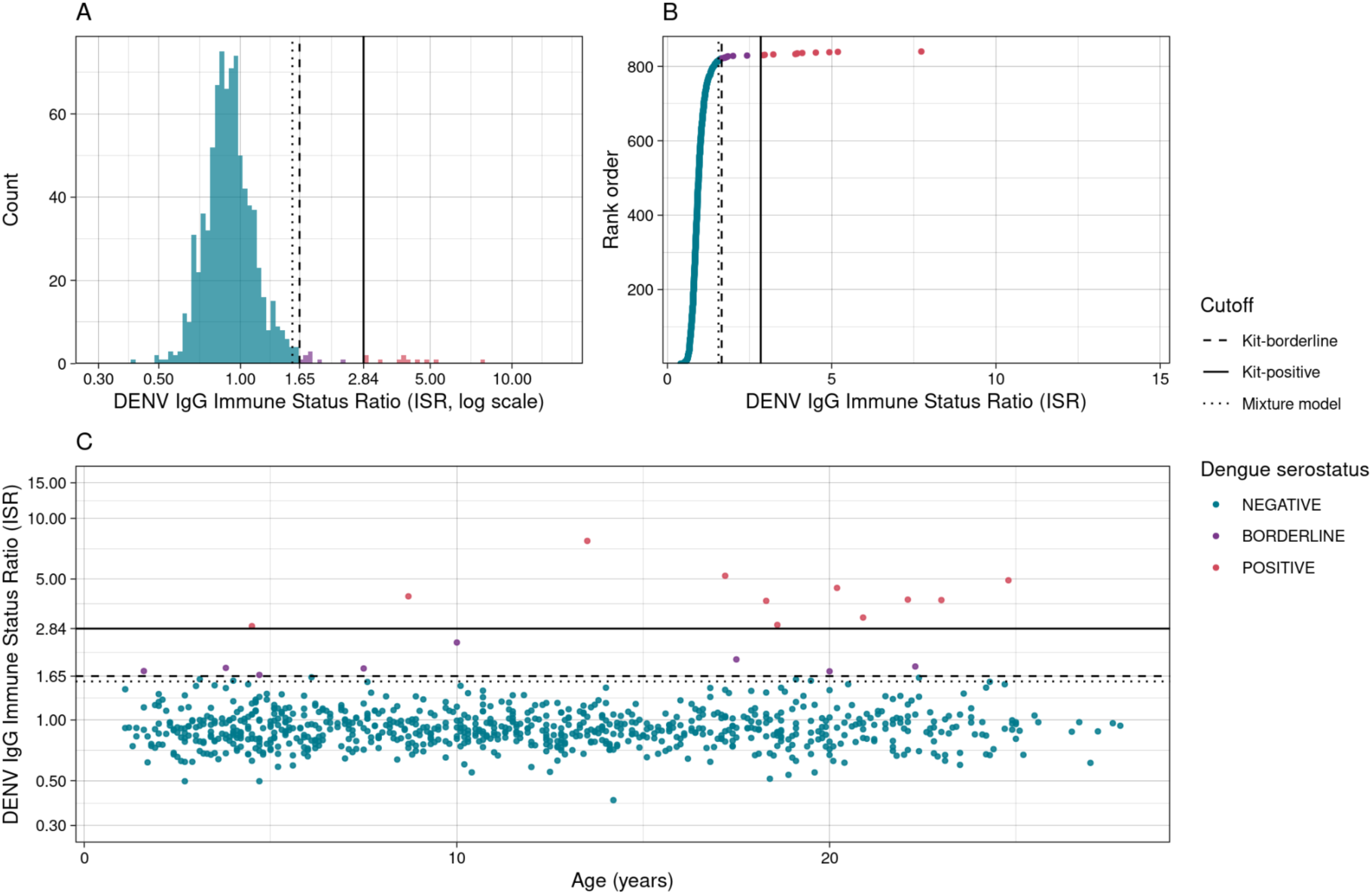
Comparison of kit-based and mixture-model cutoffs for dengue serologic classification. (A) Distribution of log-transformed DENV IgG immune status ratio (ISR) values among baseline samples from study participants (N = 840). (B) Ranked distribution of ISR values from baseline samples, with each point representing an individual observation colored by dengue serostatus classification. (C) Age-specific distribution of ISR values at baseline. Points represent individual observations colored by dengue serostatus classification, and horizontal lines indicate serologic classification thresholds. Serologic classification thresholds are shown by reference lines in each panel: the kit-positive cutoff (ISR 2.84), the kit-borderline range (ISR 1.65–2.84), and the mixture-model cutoff (ISR 1.55), estimated using a two-component Gaussian finite mixture model.

**Figure 2.**
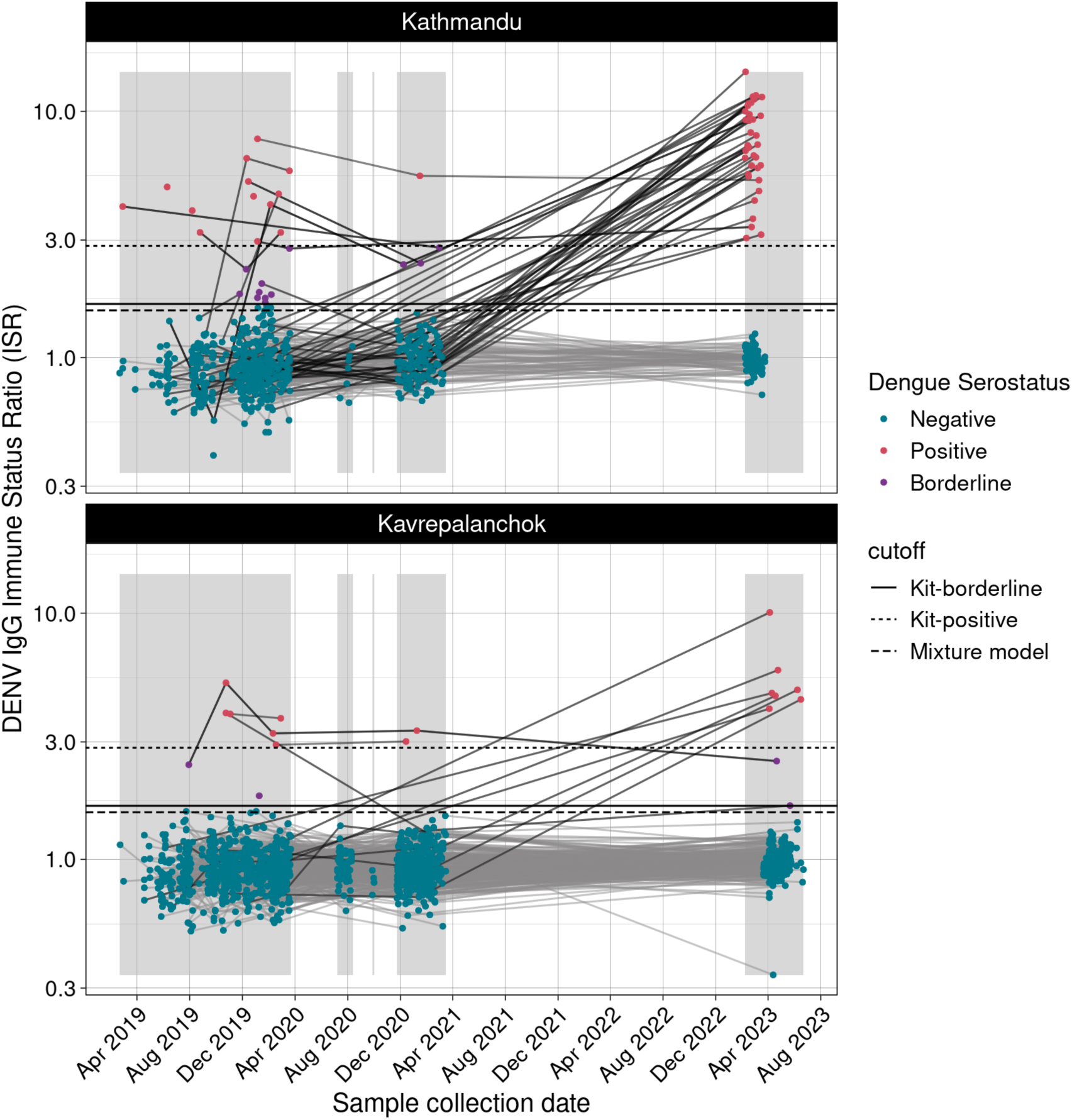
Longitudinal anti-DENV IgG responses by sampling date and study site in Kathmandu and Kavrepalanchok districts, Nepal, 2019–2023. Quantitative DENV IgG Immune Status Ratio (ISR) values from dried blood spot samples are shown by sample collection date. Points represent individual samples and are colored by dengue serostatus: red indicates seropositive samples, blue indicates seronegative samples, and purple indicates borderline results. Horizontal lines indicate classification thresholds: the solid line represents the kit-positive cutoff, the dashed line represents the kit-borderline cutoff, and the dotted line represents the mixture-model cutoff derived from a two-component Gaussian finite mixture model. Lines connect repeated measurements from the same individual across study visits; black lines connect participants with at least one seropositive result, whereas grey lines connect participants who remained seronegative throughout follow-up. Shaded grey bands indicate the participant enrollment periods for each phase of the study..

**Table 3:** Dengue seroincidence rates per 1000 person-years.

|  | Incident<br>seroconversions | Person-years | Seroincidence rate per 1000<br>person-years |
| --- | --- | --- | --- |
| Overall | 47 | 1388.8 | 33.8 (24.9-45.0) |
| **Year-wise** |  |  |  |
| 2019 | 2 | 43.4 | 46.1 (5.6-166.5) |
| 2020 | 3 | 235.1 | 12.8 (2.6-37.3) |
| 2021 | 0 | 286.8 | 0.0 (0.0-12.9) |
| 2023 | 42 | 823.5 | 51.0 (36.8-68.9) |
| **Age-group** |  |  |  |
| 0-<5 | 1 | 85.3 | 11.7 (0.3-65.3) |
| 5-<10 | 8 | 385.6 | 20.7 (9.0-40.9) |
| 10-<15 | 10 | 379.8 | 26.3 (12.6-48.4) |
| >15 | 28 | 538.1 | 52.0 (34.6-75.2) |
| **City/town** |  |  |  |
| Banepa | 2 | 224.8 | 8.9 (1.1-32.1) |
| Dhulikhel | 1 | 206.2 | 4.8 (0.1-27.0) |
| Kathmandu | 39 | 369.1 | 105.7 (75.1-144.4) |
| Panauti | 1 | 461.2 | 2.2 (0.1-12.1) |
| Panchkhal | 4 | 127.5 | 31.4 (8.5-80.3) |
| **Gender** |  |  |  |
| Female | 24 | 681.2 | 35.2 (22.6-52.4) |
| Male | 23 | 707.5 | 32.5 (20.6-48.8) |

### Factors Associated with Seroincidence of Dengue in Phase II

The strongest predictors of dengue seroconversion during follow-up were household exposure to a reported dengue case and the presence of flower pot containers with standing water. In univariate analyses (Table 4), participants living in a household with a reported dengue case had a higher risk of seroconversion than those without this exposure (RR 4.65, 95% CI 2.72– 8.00). Presence of water-filled flower basins in the household vicinity was also associated with increased risk of seroconversion (RR 2.53, 95% CI 1.28–5.74). Participants living in areas with dengue cases in the surrounding had a higher risk of seroconversion than those without (RR 4.40, 95% CI 2.55-7.72). In contrast, having window screens on all windows was associated with lower risk of seroconversion (RR 0.51, 95% CI 0.29–0.90), corresponding to a 49% lower risk. No other factors were significantly associated with seroconversion.

**Table 4:**
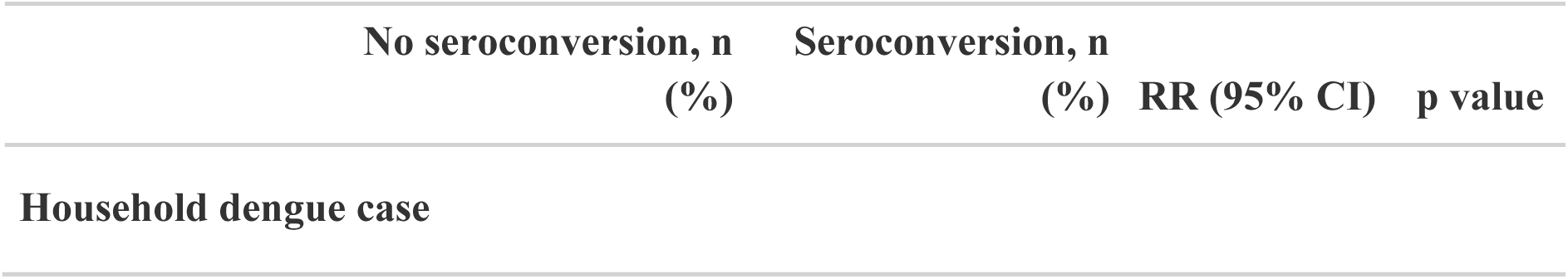
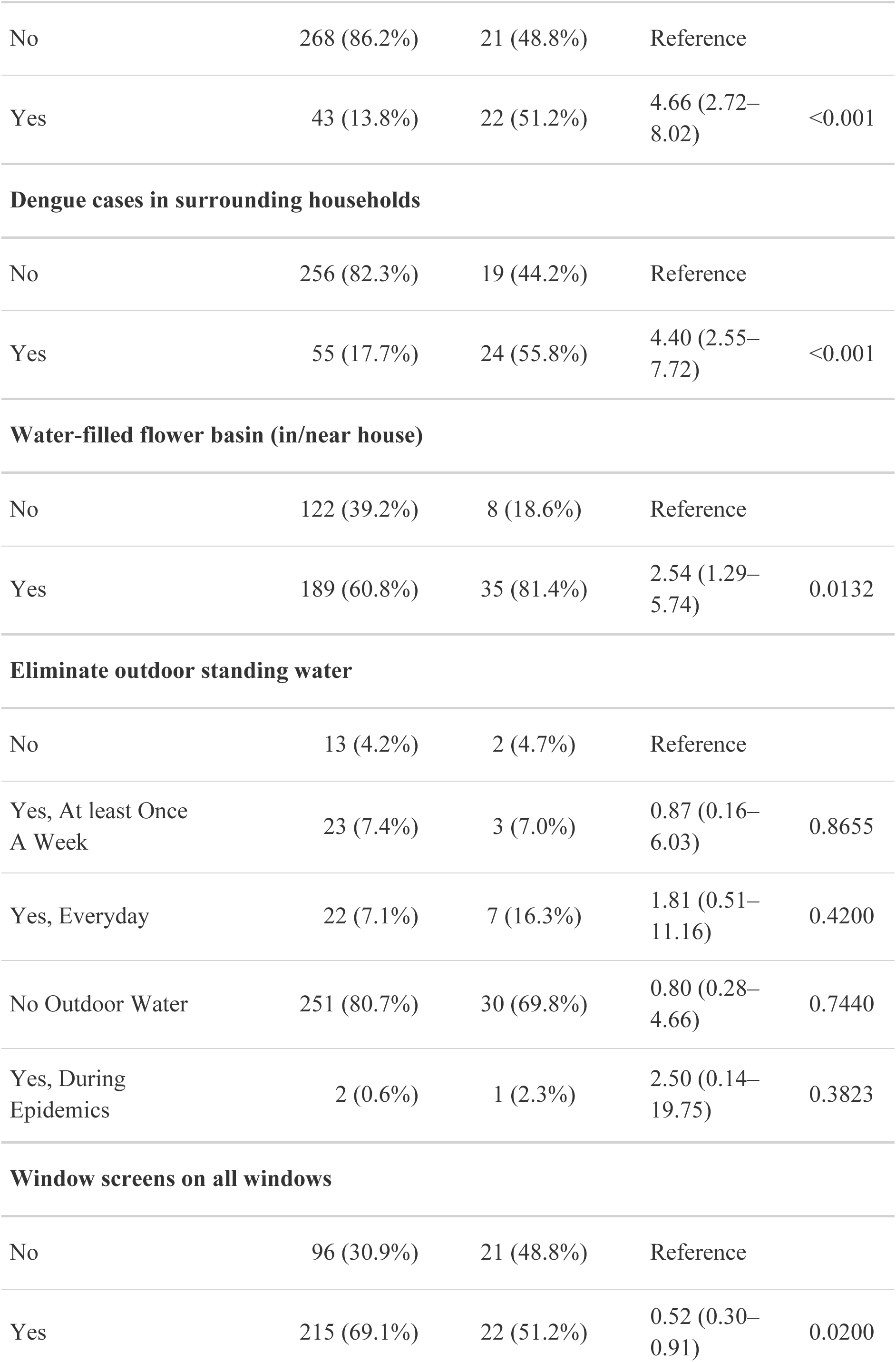
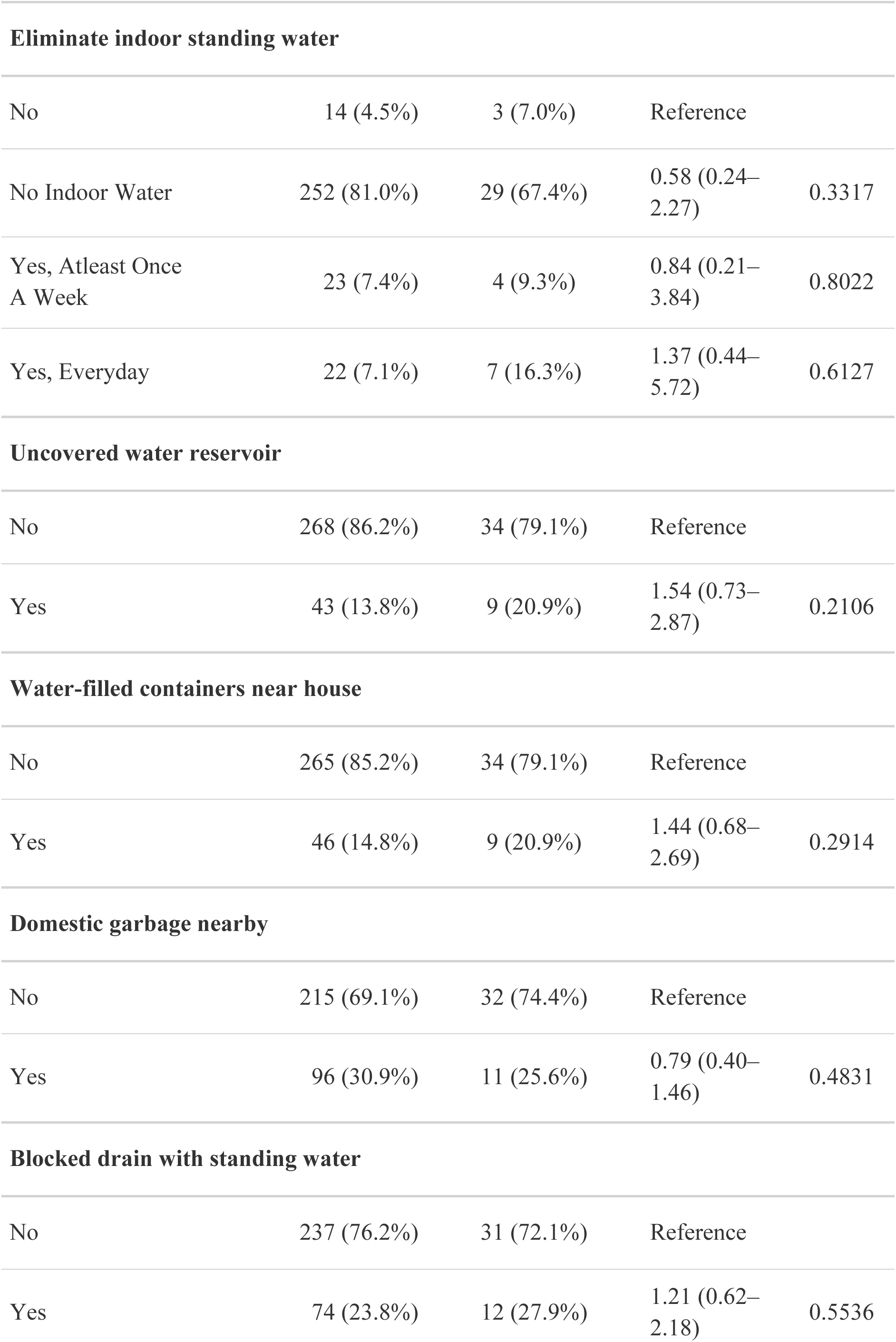
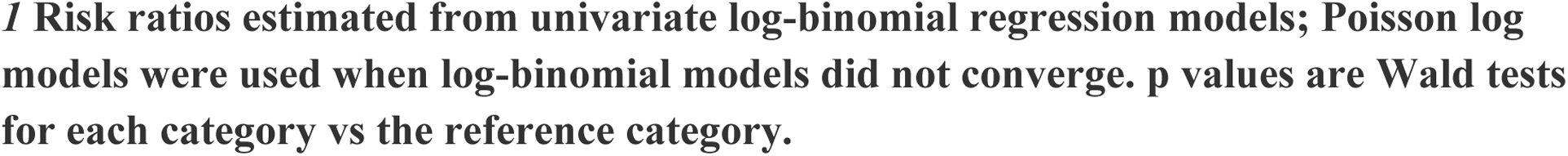
Household and environmental risk factors for dengue seroconversion during follow-up, Kathmandu and Kavrepalanchok districts, Nepal.

|  | No seroconversion, n<br>(%) | Seroconversion, n<br>(%) | RR (95% CI) | p value |
| --- | --- | --- | --- | --- |
| **Household dengue case** |  |  |  |  |
| No | 268 (86.2%) | 21 (48.8%) | Reference |  |
| Yes | 43 (13.8%) | 22 (51.2%) | 4.66 (2.72–8.02) | <0.001 |
| **Dengue cases in surrounding households** |  |  |  |  |
| No | 256 (82.3%) | 19 (44.2%) | Reference |  |
| Yes | 55 (17.7%) | 24 (55.8%) | 4.40 (2.55–7.72) | <0.001 |
| **Water-filled flower basin (in/near house)** |  |  |  |  |
| No | 122 (39.2%) | 8 (18.6%) | Reference |  |
| Yes | 189 (60.8%) | 35 (81.4%) | 2.54 (1.29–5.74) | 0.0132 |
| **Eliminate outdoor standing water** |  |  |  |  |
| No | 13 (4.2%) | 2 (4.7%) | Reference |  |
| Yes, At least Once A Week | 23 (7.4%) | 3 (7.0%) | 0.87 (0.16–6.03) | 0.8655 |
| Yes, Everyday | 22 (7.1%) | 7 (16.3%) | 1.81 (0.51–11.16) | 0.4200 |
| No Outdoor Water | 251 (80.7%) | 30 (69.8%) | 0.80 (0.28–4.66) | 0.7440 |
| Yes, During Epidemics | 2 (0.6%) | 1 (2.3%) | 2.50 (0.14–19.75) | 0.3823 |
| **Window screens on all windows** |  |  |  |  |
| No | 96 (30.9%) | 21 (48.8%) | Reference |  |
| Yes | 215 (69.1%) | 22 (51.2%) | 0.52 (0.30–0.91) | 0.0200 |
| **Eliminate indoor standing water** |  |  |  |  |
| No | 14 (4.5%) | 3 (7.0%) | Reference |  |
| No Indoor Water | 252 (81.0%) | 29 (67.4%) | 0.58 (0.24–2.27) | 0.3317 |
| Yes, Atleast Once A Week | 23 (7.4%) | 4 (9.3%) | 0.84 (0.21–3.84) | 0.8022 |
| Yes, Everyday | 22 (7.1%) | 7 (16.3%) | 1.37 (0.44–5.72) | 0.6127 |
| **Uncovered water reservoir** |  |  |  |  |
| No | 268 (86.2%) | 34 (79.1%) | Reference |  |
| Yes | 43 (13.8%) | 9 (20.9%) | 1.54 (0.73–2.87) | 0.2106 |
| **Water-filled containers near house** |  |  |  |  |
| No | 265 (85.2%) | 34 (79.1%) | Reference |  |
| Yes | 46 (14.8%) | 9 (20.9%) | 1.44 (0.68–2.69) | 0.2914 |
| **Domestic garbage nearby** |  |  |  |  |
| No | 215 (69.1%) | 32 (74.4%) | Reference |  |
| Yes | 96 (30.9%) | 11 (25.6%) | 0.79 (0.40–1.46) | 0.4831 |
| **Blocked drain with standing water** |  |  |  |  |
| No | 237 (76.2%) | 31 (72.1%) | Reference |  |
| Yes | 74 (23.8%) | 12 (27.9%) | 1.21 (0.62–2.18) | 0.5536 |

***/ Risk ratios estimated from univariate log-binomial regression models; Poisson log models were used when log-binomial models did not converge. p values are Wald tests for each category vs the reference category.***

### Geospatial analysis

The serostatus of each observation throughout the study are represented in Figure 3A. Dengue seroincidence rates are represented by the color intensity, showing high seroincidence in Kathmandu, followed by Panchkhal.

**Figure 3.**
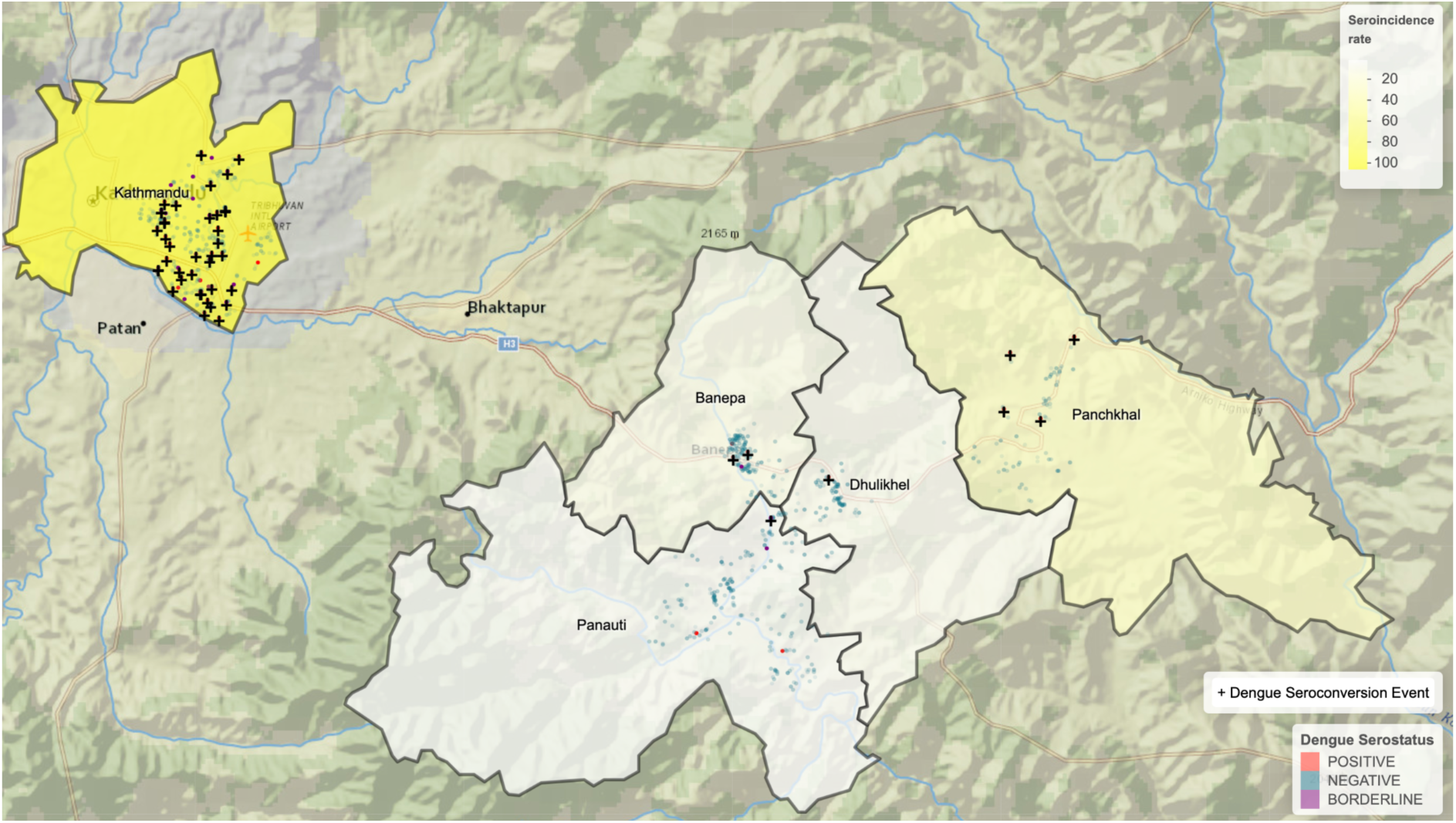
Geographic distribution of dengue seropositivity and seroincidence in Kathmandu and Kavrepalanchok districts, Nepal, 2019–2023. The map shows household locations of study participants in Kathmandu and Kavrepalanchok districts. The background shading represents the dengue seroincidence rate per 1,000 person-years estimated for each municipality. Points represent individual observations from the longitudinal serosurvey: red circles indicate dengue seropositive samples, blue circles indicate seronegative samples, and purple circles indicate borderline serologic results. Black plus symbols (+) indicate dengue seroconversion events identified during follow-up.

### Sensitivity analyses

Seroincidence rates by age, city and gender did not change substantially when using the mixture model cutoff in comparison to the kit-provided cutoff (Supplemental Table 1). Similarly, the seroprevalence by year was similar regardless of cutoff method (Supplemental Table 2).

## DISCUSSION

In this longitudinal serosurvey of 840 participants aged 0–26 years in central Nepal, we observed a marked increase in dengue seroprevalence between 2019 and 2023, particularly in urban Kathmandu where seroprevalence rose from 4.3% to 34.0%. The overall seroincidence rate was 33.8 per 1,000 person-years, with pronounced geographic heterogeneity: rates in Kathmandu (105.7 per 1,000 person-years) were nearly 50-fold higher than in Panauti (2.2 per 1,000 person-years), a municipality at similar elevation in neighboring Kavre district. Living in a household with a reported dengue case and the presence of water-filled flower basins were associated with increased risk of seroconversion, while window screens were protective. Dengue seroprevalence increased with age in both study districts, as expected given cumulative exposure over time. However, we also observed higher seroincidence rates among participants aged 15 years and older (52.0 per 1,000 person-years) compared to younger children (11.7 per 1,000 person-years among those <5 years). This pattern differs from findings in hyperendemic settings, where seroincidence is often highest among young children. In a prospective cohort in Colombo, Sri Lanka (2016–2019), dengue incidence was highest among children under 12 years (21–23 per 1,000 person-years) and lowest among adults (9.2 per 1,000 person-years) (34). Similarly, in Managua, Nicaragua (2004–2010), the majority of incident infections occurred in children under 9 years, with an overall incidence of 90.2 per 1,000 person-years (35). The higher seroincidence among older adolescents and young adults in our study may reflect the relatively recent establishment of endemic transmission in the Kathmandu Valley. In other settings where dengue was prevalent the older children would have prior immunity. Meanwhile, in a setting in high-transmission urban areas with recent introduction, both age groups are immune naive and may have different exposure such as greater mobility and occupational exposures driving the risk of dengue.

Dengue seroincidence was higher in Kathmandu, compared to Kavre. Kathmandu, the capital city of Nepal, has a population of more than 1 million and with the population density of 5,169 persons per sq. km., which could play a role in transmission of disease as well (46,47) while the population density of Kavrepalanchowk is 261 persons per sq. km. (48). The median elevation in Kathmandu is 1300 m (range 1900-2800) and the temperature ranges from (3-29)°C (36). While Kavre district neighbors Kathmandu, it has a slightly higher median elevation of 2012 m (range 1007 - 3018) and lower temperatures of (2-28)°C (36–39). Up until 2009, dengue was more prevalent in the low land regions (67-300m) of Nepal; however, in 2010 dengue fever affected highland districts of Dhading (1700m) and Kathmandu (40,41). Since then the cases are rising and have been documented to all the 77 districts as of 2022 (42). Rising temperatures and rainfall is a possible explanation of the rise of dengue transmission at higher elevations in Nepal. The increased temperature from (26 - 28)°C to 30 °C decreases the extrinsic incubation period of dengue virus in mosquitoes promoting the viral transmission by being effective (43–45). Climate modeling using General Circulation Models (GCMs)— computational models that simulate atmospheric and oceanic dynamics—has projected that temperatures in Nepal will increase between 0.5°C and 2.0°C by 2030 and 3.0°C to 6.3°C by the 2090s. These projections suggest that conditions suitable for Aedes mosquito survival and dengue virus transmission will continue to expand to higher altitudes (46).

In our study, individuals who lived in dwellings where there is presence of a water-filled flower basin had 2.5x higher risk of dengue incidence compared to individuals living in households without a basin. In Nepal, open flower basins are commonly used as a decoration. These basins are filled with standing water and flowers and may act as a breeding site for mosquitoes (47,48). In a few studies conducted in Malaysia, flower basins/pots were identified as the major mosquito breeding sites (49,50). Public health officials could use this information to educate the public about changing water in decorative basins frequently or treating it with insecticide to limit aedes mosquito breeding (51). The presence of other potential breeding sites for aedes mosquitoes such as blocked drain, nearby domestic garbage, uncovered water container, tires and presence of containers/tires/tins near houses were also associated with an elevated risk of dengue, however confidence intervals for incidence ratios included 0. With a sample size of 354, we were underpowered to detect effect sizes less than 2.

The strongest risk factor for dengue seroconversion was residing in the house with seroprevalent cases, which was associated with a nearly five times higher seroconversion rate. This suggests that a significant proportion of dengue transmission is occurring at the household level either because of household-level transmission or shared risk factors. A cohort study conducted in Kamphaeng Phet province in Northern Thailand (2015-2021) showed the household members had 1.26 higher odds of being seropositive in the household having a seropositive case (52). The existence of household clustering of cases has been described in several studies (53,54). Together, these findings suggest it is important to perform the indoor spraying and educational campaign about the risk factors of dengue at the household level and ways to prevent it.

The results of this study should be interpreted in light of several limitations. First, only 42% (354/840) of the original cohort was available to enroll in Phase II, when household and environmental risk factors were assessed. Differential loss to follow-up may have introduced selection bias; if individuals at higher or lower risk of dengue were more likely to drop out, this could have biased seroincidence estimates and attenuated associations with risk factors. Second, because risk factor data were collected only in Phase II, we cannot establish temporal relationships between environmental exposures and seroconversion events that occurred earlier in the study. Third, we limited enrollment to participants aged 0–26 years, and therefore we cannot determine whether seroincidence, seroprevalence, or risk factors differ among older age groups. Fourth, the IgG ELISA used in this study cannot distinguish between dengue serotypes or differentiate primary from secondary infections, which have different clinical implications. Finally, although the InBios DENV Detect™ assay has high reported specificity (97.7%), cross-reactivity with other flaviviruses endemic to Nepal, such as Japanese encephalitis virus, cannot be entirely excluded and may have resulted in some misclassification of serostatus.

Despite these limitations, to our knowledge this is the first population-based longitudinal serosurvey of dengue in Nepal. The longitudinal design allowed direct estimation of seroincidence rates during a period of rapidly increasing dengue transmission in the Kathmandu Valley, and the community-based sampling provides estimates that are more representative of the true infection burden than hospital-based surveillance alone.

With seroincidence rates in urban Kathmandu approaching those of hyperendemic settings in South and Southeast Asia, these findings demonstrate that intense dengue transmission is now established in Nepal’s densely populated capital—a high-altitude area previously considered at low risk. The substantially lower seroincidence in neighboring rural districts of Kavrepalanchok, despite similar elevations, suggests that urbanization and population density may be stronger determinants of transmission than altitude alone. These data underscore the need for enhanced surveillance systems that integrate periodic serosurveys with case-based reporting to monitor transmission intensity and inform future vaccine introduction decisions. In the interim, household-level interventions targeting water-filled decorative containers and indoor residual spraying in households with confirmed cases offer actionable strategies to reduce transmission as dengue becomes established in Nepal’s urban highland areas.

## Data Availability

All data produced in the present work are contained in the manuscript

## LIST OF ABBREVIATIONS

ELISA: Enzyme-Linked Immunosorbent Assay
ISR: Immune Status Ratio
IgG: Immunoglobulin G
DENV: Dengue Virus
RR: Risk Ratio
CI: Confidence Interval
PBS: Phosphate Buffer Saline
DBS: Dried Blood Spot Samples
DENRA: Dengue-Derived Recombinant Antigen
NCA: Normal Cell Antigen
GCMs: General Circulation Models
IQR: Interquartile Range

## DECLARATIONS

## Ethics approval and consent to participate

The study and protocol was approved by the Nepal Health Research Council (ref no., 1544), Ethics Review Board of Dhulikhel Hospital and Stanford University. Written informed consent form was obtained from the participants above 18 years old. Written assent form was obtained from the participants of age 15 and 17. Additionally, for the minors under the age of 18, legal guardians or parental consent were obtained.

## Consent for Publication

Not applicable

## Availability of Data and Material

Not applicable

## Authors contributions

AS: Data Acquisition, Analysis, Interpretation, Original Draft, Manuscript Revision

MT: Laboratory study, Manuscript Review

SS: Laboratory study, Manuscript Review

ST: Laboratory study, Data Acquisition, Manuscript Review

UR: Laboratory study, Data Acquisition, Manuscript Review

NK: Laboratory study, Data Acquisition, Manuscript Review

SBS: Laboratory study, Manuscript Review

SRN: Laboratory study, Manuscript Review

JRA: Conceptualization, Overall Study Design, Manuscript Review

RS: Conceptualization, Overall Study Design, Data Acquisition, Manuscript Review

KA: Conceptualization, Overall Study Design, Data Analysis, Interpretation, Manuscript Revision

DT: Conceptualization, Overall Study Design, Data acquisition, Data Analysis, Interpretation, Original Draft, Manuscript review

## Acknowledgements

We gratefully acknowledge each individual who participated in this study and all the authors for their valuable contributions.

---

**Supplemental Table 1:**
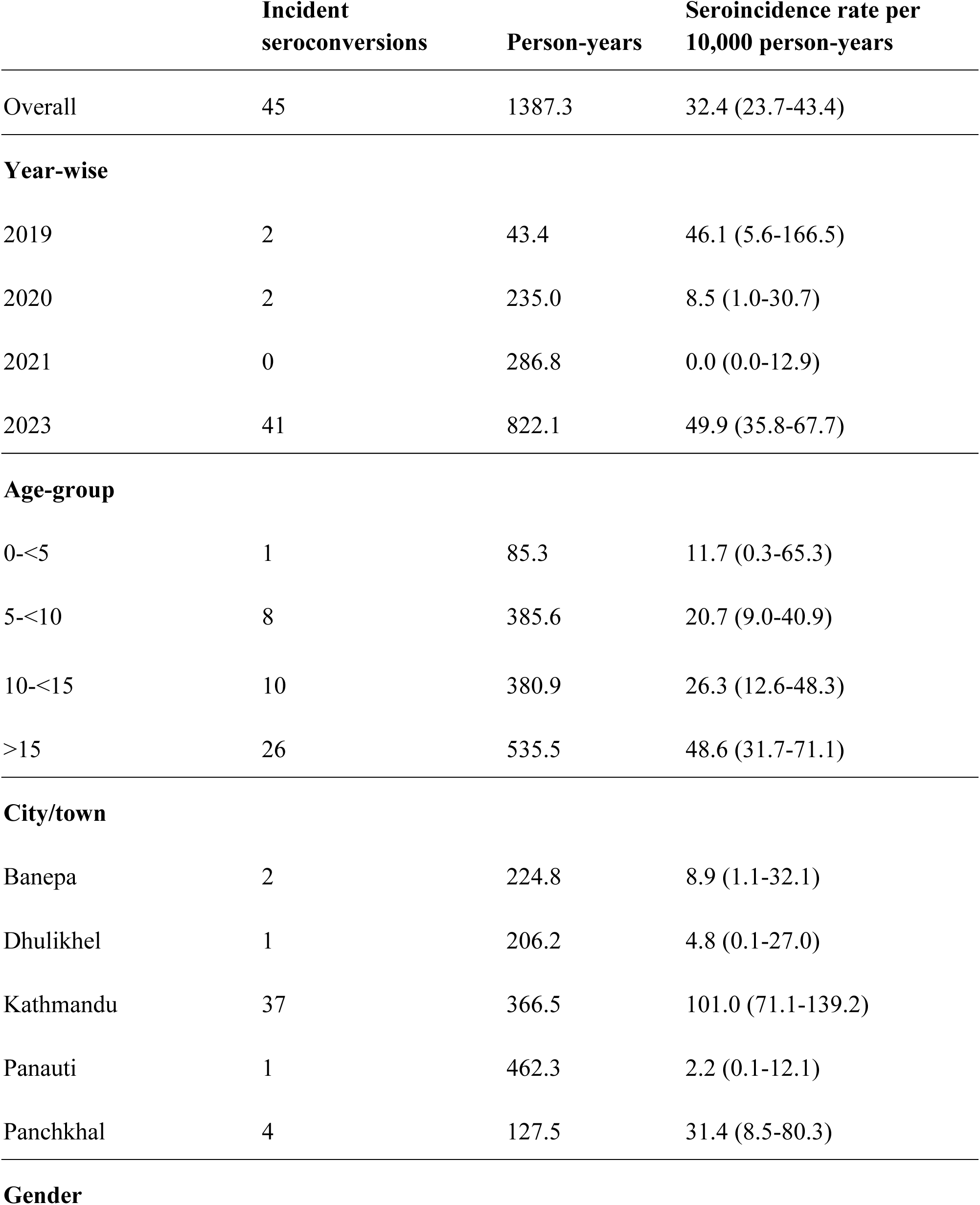
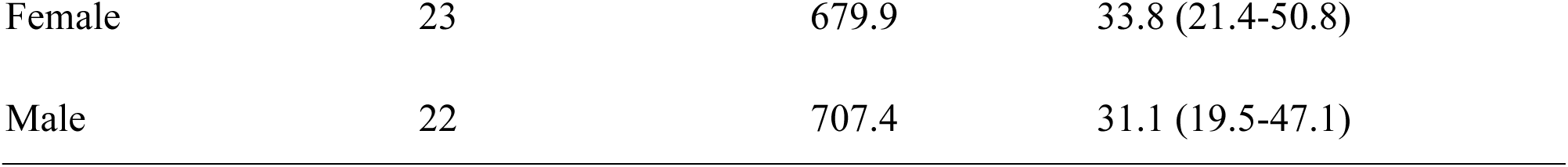
Sensitivity analysis of dengue seroincidence rates per 1000 person-years using the mixture model cutoff.

**Supplemental Table 2:**
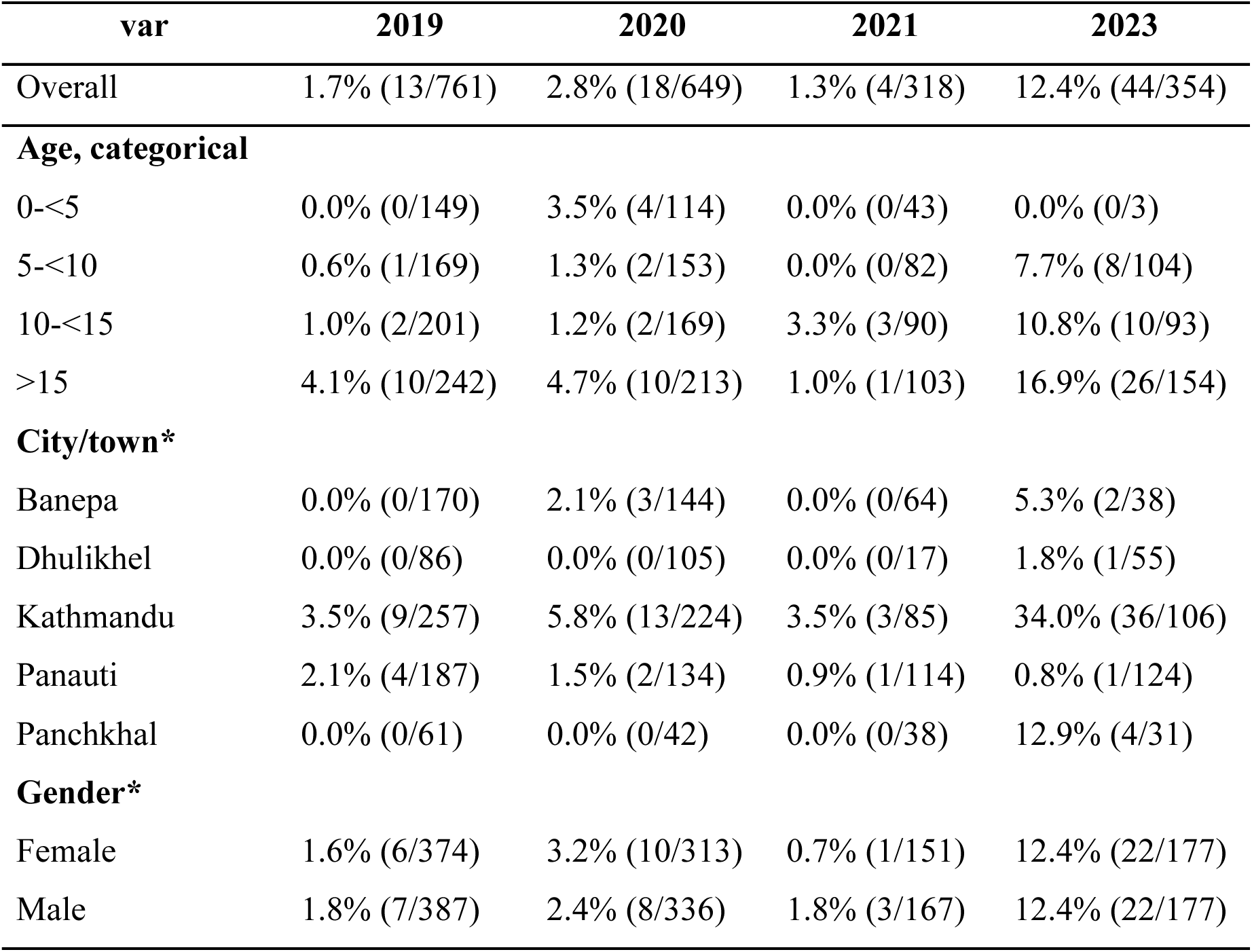
Sensitivity analysis of dengue seroprevalence by year using the mixture model cutoff.

